# Performance characteristics of the SARS-CoV-2 Isothermal Molecular Assay for point-of-care molecular diagnosis of SARS-CoV-2

**DOI:** 10.1101/2025.03.29.25324880

**Authors:** Chika Kingsley Onwuamah, Joseph Ojonugwa Shaibu, Olufemi Samuel Amoo, Rahaman A. Ahmed, Bowofoluwa Sharon Abimbola, Rosemary Ajuma Audu, Babatunde Lawal Salako

## Abstract

**Background:** Access to diagnostics was a significant challenge during the early stages of the COVID-19 pandemic, as laboratory reagents, supplies, and consumables were in short supply and high demand. Central laboratories, equipped with advanced real-time PCR equipment and highly skilled staff, worked long hours to produce test results for travellers, hospital patients, and the general public. We developed a SARS-CoV-2 molecular diagnostic assay suitable for point-of-care testing to address this diagnostic challenge.

**Methods:** Recombinase amplification coupled with fluorescent detection was chosen as the technology, as it was freely available. Using a mobile fluorescent detection device ensures foolproof results interpretation, point-of-care testing and minimal footprint in facilities. Designed as a one-step reverse transcriptase real-time isothermal assay utilising the FAM and ROX channels. Specimens were tested using commercially available real-time PCR assays to evaluate the performance characteristics of the assay.

**Results:** The SARS-CoV-2 Isothermal Molecular Assay (SIMA) detects the SARS-CoV-2 ORF1ab gene and a human gene to differentiate poor specimen collection. SIMA runs at 40°C for 36 minutes, with the first six minutes dedicated to reverse transcription. It has a detection limit of 555 RNA copies/mL. Highly positive specimens are detected within three minutes, but the 30-minute run time allows for detecting low-positive Specimens. Sensitivity was 95%, with 100% specificity, while precision ranged from 4% to 6%

**Conclusions:** We report the performance characteristics of SIMA, the first molecular diagnostics developed in Nigeria for SARS-CoV-2 point-of-care diagnosis. Its performance and utility give hope for rapid diagnosis and treatment in resource-constrained settings.

## Introduction

The COVID-19 pandemic has had a profound impact globally. Still, preparedness systems and existing human and infrastructural capacities helped many nations and institutions survive, thrive, and rise to the occasion, as demonstrated by the Nigerian Institute of Medical Research in Lagos (NIMR), Nigeria (1). NIMR performed the first partial sequencing of SARS-CoV-2 in Nigeria to confirm the index case and offered free testing to the public in Lagos, Nigeria’s megacity with over 20 million residents (2). To allay public panic, we described the pandemic among infected individuals presenting at our facilities, determined that COVID-19 spread patterns were unrelated to malaria co-infections, and confirmed that blood transfusion was safe (3-6). We collaborated across Africa to conduct genomic surveillance for SARS-CoV-2, publishing a protocol for Sanger sequencing of the virus, as this sequencing platform was widely available in Africa prior to the COVID-19 pandemic (7-11). Health workers were reassured that routine infection control practices still worked and that the vaccine they received as a priority cohort was effective (12-13). However, diagnostics was a significant challenge at the time, including collecting the specimens for testing. After many health workers became infected and quarantined, we brought a do-it-yourself specimen collection protocol (14). To address the diagnostic challenge, we mentored emerging testing laboratories and published our experience with five commercially available diagnostic assays, helping laboratories that we could not reach physically (15). We further developed molecular diagnostic assays, including one for routine PCR systems and another for near-point-of-care use to decentralise testing (16). This manuscript describes the performance evaluation and characteristics of the near-point-of-care assay, known as the SARS-CoV-2 Isothermal Molecular Assay (SIMA).

Diagnostics are critical to ensure patients are treated for the right ailment the first time, reducing hospital stay and costs, and can be optimised to direct appropriate treatment in an era of increasing drug resistance (15). The appropriate use of diagnostic tools preserves available treatment options and monitors disease patterns, which helps implement effective health interventions. Health and laboratory personnel were severely strained early in the COVID-19 pandemic, particularly during the global lockdown (1). Laboratories had to work long, hard hours to churn out SARS-CoV-2 test results, whether for travellers, hospital patients or the general populace. Centralised tertiary facilities with highly skilled staff and advanced, real-time PCR equipment were performing the diagnostic testing (15). However, access to laboratory reagents, supplies and consumables was limited and stretched due to the global lockdown. In Nigeria, after the death of some clinical staff who attended to COVID-19 patients unknowingly and without adequate personal protective equipment, patients were mainly unattended until their COVID-19 status was known. The laboratory test turnaround time in Nigeria during the pandemic’s peak stretched from two weeks to approximately two months, depending on the scenario and location (17). Thus, patients could deteriorate while awaiting COVID-19 test results.

Amidst these challenging times, we have developed a diagnostic assay that reliably detects infected individuals, particularly those with a sufficient viral load to infect others. Most importantly, we sought to utilise alternative diagnostic technologies whose availability would not have been affected by the fact that demand had outstripped the supply chain for routine diagnostic technologies. A key benefit was to ensure it was a near-point-of-care assay, enabling small and remote health facilities to perform testing and improve patient care while alleviating the heavy burden on central laboratories. Consequently, it was expedient for the assay system to be fool-proof, such that anyone without the high expertise required for real-time PCR could be quickly trained to perform the testing qualitatively.

Recombinase Amplification is an isothermal molecular amplification technique that has proven highly sensitive and specific, eliminating the need for expensive thermal cycling equipment (18). It has been used several times to detect pathogens, polymorphisms, etc. (19). Recombinase amplification runs typically at 39-42□, giving results in 5-30 minutes, and test results can be visualised in different means (20). Amplification can even be incubated using body heat (20). This isothermal amplification technique is well-suited for point-of-care applications, particularly in resource-constrained settings (20). Its potential for point-of-care use was exploited to develop a molecular diagnostic assay for SARS-CoV-2, which is detected using fluorescence in a portal device to minimise subjective interpretation, even when used in remote locations by non-technical personnel trained to perform the workflow, e.g., Community Health Extension Workers.

The SARS-CoV-2 Isothermal Molecular Assay (SIMA) is a molecular test that detects the virus causing COVID-19. It is intended to detect infected persons, especially ones with a high viral load that makes them infectious to others (Ct values ≤ 25). The advantages of recombinase isothermal amplification include its robustness and speed, with detection possible in minutes using multiple options (20). It features a straightforward workflow, and amplification is possible at low temperatures (20 °C). Its low-cost detection equipment and test reagents have proven to be sensitive and specific. More importantly, low-technical staff can be trained to perform SIMA, and it can be used in any cadre of healthcare facilities, even powered all day by a small battery pack.

We report the in-house performance evaluation of the assay, developed during the COVID-19 global lockdown, to enable any health facility in Nigeria to perform SARS-CoV-2 diagnosis reliably and efficiently.

## Methodology

### Kit design and formulation

SIMA currently detects one SARS-CoV-2 target, the ORF1ab gene. It is a one-step reverse transcriptase real-time isothermal assay that detects SARS-CoV-2 ORF1ab on FAM and a human housekeeping gene on ROX. The assay’s detection limit is influenced by the reverse transcriptase enzyme’s ability to transcribe SARS-CoV-2 RNA into complementary DNA at a relatively low temperature. SIMA runs at 40 °C for 36 minutes, with the first six minutes dedicated to the reverse transcription of the SARS-CoV-2 RNA specimen without shaking or magnetic stirring. The six-minute reverse transcriptase step converts SARS-CoV-2 RNA to complementary DNA. After that, amplification continues for a maximum of 30 minutes to enable the amplification of low-viral-load specimens. High positive specimens are detected as early as three to nine minutes after assay initiation. SIMA testing is more efficient in batch analysis comprising 1-14 unknown specimens, one negative and one positive control. The negative and positive controls must meet the thresholds below for the assay to be valid.

| Control | Target | Time to rising (Sec) |  |  | Last Fluorescent Read (Mv) |  |  |
| --- | --- | --- | --- | --- | --- | --- | --- |
|  |  | Average | Min | Max | Average | Min | Max |
| Negative | SARS (FAM) | - | - | - | 270.9 | 211.3 | 800.0 |
|  | IC (ROX) | 725.6 | 637.0 | 824.0 | 2659.7 | 2429.1 | 2970.9 |
| Positive | SARS (FAM) | 609.4 | 534.0 | 692.0 | 3783.3 | 1500.0 | 5000.0 |
|  | IC (ROX) | 728.8 | 652.0 | 848.0 | 2624.3 | 1500.0 | 5000.0 |

If testing more specimens on the same day, subsequent batch testing does not need to involve controls, as daily testing of controls is sufficient. The T8-ISO and T16-ISO devices (Axxin, Australia) were used to detect amplification in this evaluation. The negative control must have FAM fluorescence below 800 and ROX fluorescence above 1000, while the positive control must have FAM fluorescence above 2000 and ROX fluorescence above 1500. Thus, the negative control must be displayed as “Negative” on the equipment, as it contains a synthetic fragment of the human gene, detected as an internal control. The positive control MUST be “Positive” as it contains synthetic gene fragments of the SARS-CoV-2 ORF1ab gene and the human gene detected as an internal control.

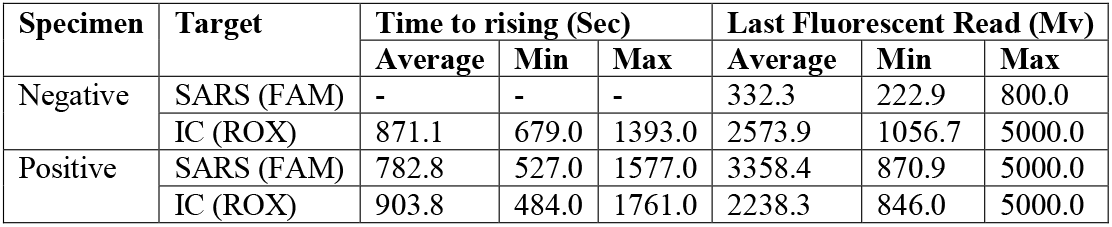

| Specimen | Target | Time to rising (Sec) |  |  | Last Fluorescent Read (Mv) |  |  |
| --- | --- | --- | --- | --- | --- | --- | --- |
|  |  | Average | Min | Max | Average | Min | Max |
| Negative | SARS (FAM) | - | - | - | 332.3 | 222.9 | 800.0 |
|  | IC (ROX) | 871.1 | 679.0 | 1393.0 | 2573.9 | 1056.7 | 5000.0 |
| Positive | SARS (FAM) | 782.8 | 527.0 | 1577.0 | 3358.4 | 870.9 | 5000.0 |
|  | IC (ROX) | 903.8 | 484.0 | 1761.0 | 2238.3 | 846.0 | 5000.0 |

A negative specimen must have FAM fluorescence below 800, with the internal control detected at ROX fluorescence above 1000. A positive specimen must have FAM fluorescence above 800; however, its internal control may not be detected due to competition or low copy numbers of the human gene. If the internal control of a positive specimen is detected, the ROX fluorescence is usually above 800. The equipment must display a negative specimen as “Negative” and a positive Specimen as “Positive”. If these parameters are not realised, then repeat testing, preferably using a fresh specimen, is recommended.

SIMA was designed for isothermal molecular amplification and fluorescent detection. It is capable of testing 1-16 reactions per run, simultaneously detecting fluorescence on both the FAM and ROX channels. Hence, the assay uses probes and minimal footprint fluorescent devices from Axxin (Australia) or Optigene (UK) can be used as detection devices. These devices are ordered with a field carrying sturdy case, that can accommodate the pippetors, tips and battery pack where necessary. Thresholds for accepting the positive and negative controls as valid and independently determining the status of the unknown specimens are configured into the detection device to ensure appropriate interpretation and preclude subjectivity. SIMA uses specialised probes, it can also be detected using routine real-time PCR equipment with the correct settings, including turning off thermal cycling and top-lid heating.

SIMA was validated in-house according to Rabenau et al. and then externally validated by other independent facilities (21). Positive specimens were selected based on at least one log10 value exceeding the assay limit of detection, while low-positive specimens were those with values between the assay limit of detection and up to one log10 value above it (21).

### SIMA Testing Procedure

Clinical specimens were collected in a VTM. RNA extraction was performed using commercially available manual or automated extraction kits. The facility used the manual QIAamp Viral RNA mini kit (Qiagen) and T142H Viral RNA and DNA automated kit on a Libex machine (Xi’an Tianlong). The SIMA testing procedure is outlined briefly below. Thaw all reagents to room temperature and vortex Reagents A and C to ensure thorough mixing. Then, briefly spin down Reagents A to C. Reagent B is an enzyme mixture that should not be vortexed and must be kept cold while in use. The table below guides the preparation of the master mix using Reagents A and B, depending on the number of tests. Include one negative and one positive control per run.

| Tests<br>→ | Volumes in microliters (µL) |  |  |  |  |  |  |  |  |  |  |  |  |  |  |  |
| --- | --- | --- | --- | --- | --- | --- | --- | --- | --- | --- | --- | --- | --- | --- | --- | --- |
|  | 1X | 2X | 3X | 4X | 5X | 6X | 7X | 8X | 9X | 10X | 11X | 12X | 13X | 14X | 15X | 16X |
| Rgt A | 42.8 | 85.6 | 128.4 | 171.2 | 214.0 | 256.8 | 299.6 | 342.4 | 385.2 | 428.0 | 470.8 | 513.6 | 556.4 | 599.2 | 642.0 | 684.8 |
| Rgt B | 3.9 | 7.8 | 11.7 | 15.6 | 19.5 | 23.4 | 27.3 | 31.2 | 35.1 | 39.0 | 42.9 | 46.8 | 50.7 | 54.6 | 58.5 | 62.4 |
| Each tube gets 44.5 µL |  |  |  |  |  |  |  |  |  |  |  |  |  |  |  |  |

Vortex the master mix tube for 15 seconds to mix, then spin it down briefly. Add 44.5 µL of the master mix to each lyophilised reagent tube containing a magnetic ball. Seal with the strip cover and mix vigorously to reconstitute the lyophilised reagents. Ensure that all powder has dissolved and briefly spin down the tube. Label the 8-tube strips on the side with the serial number as on the run sheet. Open the strip cover and add 2.5 µL of Reagent C to the cap of each tube. Add 3.0 µL of controls/specimens to be tested into the designated tube and mix. Cover the tube strips with their covers containing reagent D and verify that all have reagent D. Start the test type protocol (SARS-IC Test type) on the isothermal instrument. Enter the details and positions of the controls and specimens, and allow the Isothermal instrument to reach the run temperature (40D). Now, spin down reagent D into the reagent and specimen mixture, and put the strip into the isothermal instrument to commence testing and watch the specimen labels. Start the test workflow.

### Standardising the Reference Assays

An RNA standard curve was determined for the two real-time PCR assays used as reference assays, the BGI single-target assay and the Genefinder three-target assay. A specimen with a high Ct value was quantified using the Qubit RNA HS assay and converted to RNA copies per microliter. The RNA standard curve consisted of six specimens in a ten-fold serial dilution. The logarithmic regression equation for each assay was used to estimate the RNA concentration per specimen based on its Ct value in the assay. All tests were performed according to the manufacturer’s instructions on the QuantStudio 3 real-time PCR system. The GeneFinder kit’s internal control was not detected at the time as emphasise was on performing both tests on the same equipment.

### Sensitivity and Specificity Evaluation

SIMA was evaluated using RNA specimens from individuals who were positive for SARS-CoV-2 at NIMR’s testing facility, including those with low-positive results. SIMA was further tested with specimens negative for SARS-CoV-2 on the reference assays, including viral transport medium (VTM) as blank controls and clinical specimens. The RNA specimens were extracted and retested on the reference assay within 24–48 hours of initial evaluation with SIMA. The data was used to calculate SIMA’s performance characteristics using the MedCalc Diagnostic Test Evaluation Calculator (22).

### Limit of Detection and Linearity Analysis

The detection limit for SIMA was evaluated using a serially diluted high-positive clinical specimen. The high-positive specimen was tested with the BGI ORF1ab assay on the QuantStudio 3 real-time PCR system. It was subsequently quantified with Qubit RNA HS assay and converted to RNA copy numbers using the formula below.

The specimen’s neat concentration was diluted to 1.0 × 10^9^ using 1X Tris-EDTA (TE) buffer. 450 µL of 1X TE buffer was added to clean labelled tubes in readiness for the specimen serial dilution. 50 µL of each preceding specimen concentration was added to 450 µL of TE buffer in the labelled tube for the subsequent dilution. The mixture was gently vortexed for 10 seconds and then spun down before 50 µL was taken for subsequent dilution. This procedure was repeated until the specimen was serially diluted from 10^9^ to 10^3^ (1 copy/µL). Each concentration was further aliquoted into single-use tubes (55 µL) to facilitate single use and minimise specimen deterioration due to freeze-thaw cycles. The single-use volume 55 allowed for testing ten replicate tests using 3–5 µL of specimen per time.

### Precision

The intra-assay precision was determined by testing ten specimens in triplicate within the same run, including specimens with low and high positive results. The Coefficient of Variation (CV; %) was determined for each specimen for the “time to rise” (≥ 700 fluorescence used as threshold) and for the last read fluorescence value. The inter-assay precision was determined in two ways. First, four specimens were tested once daily over three different days. Secondly, three specimens were tested twice on the same day but in different runs. The Coefficient of Variation (CV; %) was determined for each specimen for the “time to rise” (≥ 700 fluorescence used as threshold) and for the last read fluorescence value.

Broad consent for diagnostic testing and sequencing was obtained from patients whose specimens were used for this evaluation and validation.

## RESULTS

As stipulated above, an RNA standard curve was first determined for both reference assays to relate the Ct values obtained from the different assays. A comparison of BGI and Genefinder standard curves for the SARS-CoV-2 RdRp and ORF1ab targets is shown in Figure 2.

**Figure 1:**
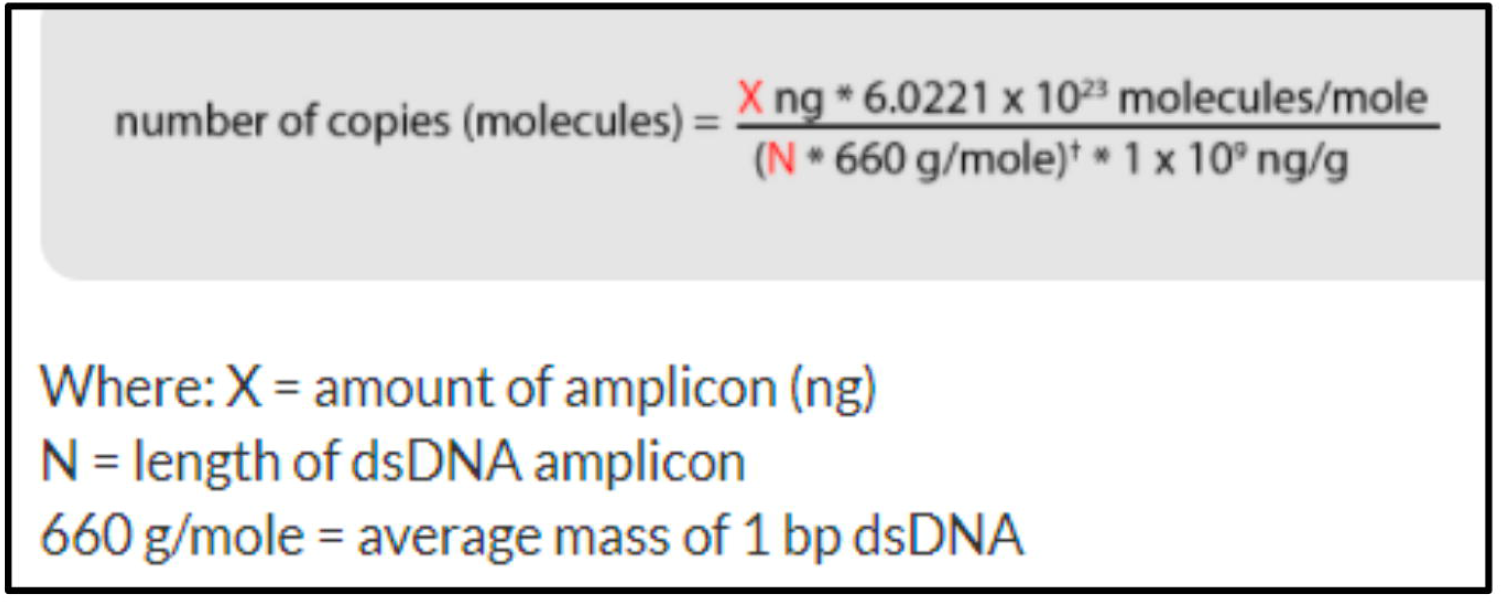
Formula for calculating estimated RNA copy numbers

**Figure 2:**
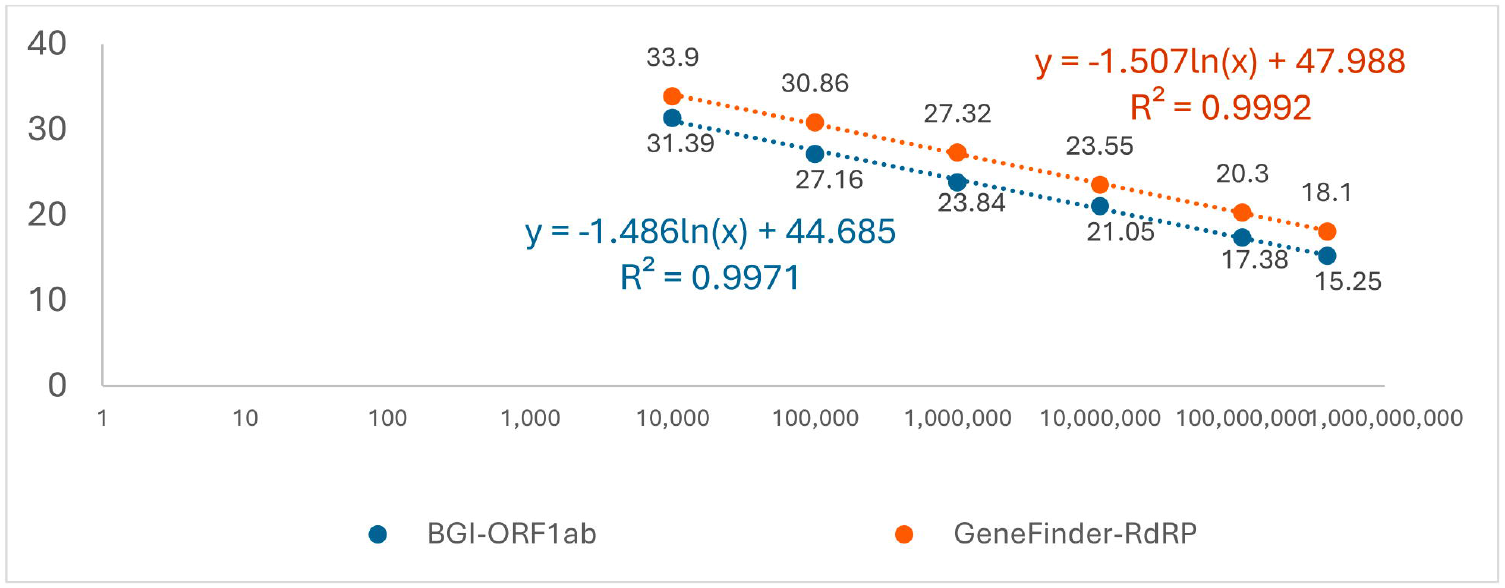
Comparison of BGI and Genefinder Ct Values

**Figure 3:**
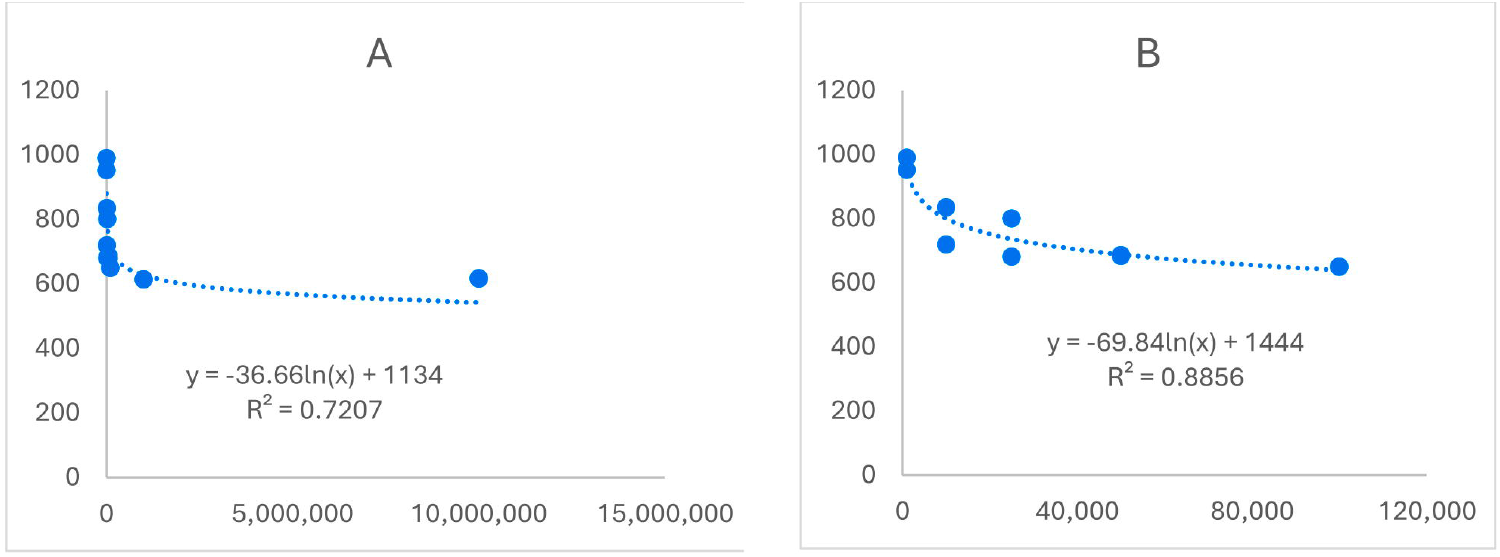
RNA Concentration vs Time to rise: A) 10^3^ - 10^7^ RNA copies/µL; B) 10^3^ - 10^5^ RNA copies/µL

### Sensitivity and Specificity

SIMA was evaluated with 91 RNA specimens from people who were positive for SARS-CoV-2. The RNA specimens used were extracted and retested on the reference assay within 24–48 hours of evaluation using SIMA. The 91 specimens comprised both low-positive specimens, with Ct values from 13.4 to 33.4.

| Specimen ID | qPCR Ct value | RNA Conc /mL | SIMA Status |
| --- | --- | --- | --- |
| S1 | 13.4 | 1,372,231.3 | Detected |
| S2 | 15.2 | 346,830.0 | Detected |
| S3 | 15.2 | 346,830.0 | Detected |
| S4 | 15.5 | 281,440.5 | Detected |
| S5 | 16.7 | 122,030.0 | Detected |
| S6 | 16.9 | 106,164.6 | Detected |
| S7 | 16.9 | 106,164.6 | Detected |
| S8 | 17.2 | 108,546.8 | Detected |
| S9 | 17.2 | 86,148.9 | Detected |
| S10 | 17.6 | 81,821.8 | Detected |
| S11 | 17.8 | 56,727.0 | Detected |
| S12 | 18.0 | 49,351.8 | Detected |
| S13 | 18.2 | 42,935.5 | Detected |
| S14 | 18.2 | 42,935.5 | Detected |
| S15 | 18.5 | 44,352.1 | Detected |
| S16 | 19.6 | 16,196.1 | Detected |
| S17 | 19.7 | 15,106.6 | Detected |
| S18 | 19.8 | 18,742.4 | Detected |
| S19 | 20.0 | 16,163.2 | Detected |
| S20 | 20.2 | 10,664.7 | Detected |
| S21 | 20.3 | 9,947.3 | Detected |
| S22 | 20.4 | 12,771.4 | Detected |
| S23 | 20.4 | 12,348.8 | Detected |
| S24 | 20.9 | 6,550.1 | Detected |
| S25 | 21.0 | 6,109.5 | Detected |
| S26 | 21.7 | 5,079.8 | Detected |
| S27 | 22.1 | 2,840.0 | Detected |
| S28 | 22.6 | 2,005.0 | Detected |
| S29 | 22.6 | 2,005.0 | Detected |
| S30 | 22.7 | 2,591.7 | Detected |
| S31 | 22.8 | 1,744.3 | Not Detected |
| S32 | 23.2 | 1,320.2 | Detected |
| S33 | 23.3 | 1,231.4 | Detected |
| S34 | 23.7 | 932.0 | Detected |
| S35 | 23.9 | 810.9 | Not Detected |
| S36 | 24.5 | 792.9 | Detected |
| S37 | 24.6 | 498.0 | Detected |
| S38 | 24.7 | 464.5 | Detected |
| S39 | 25.0 | 555.0 | Detected |
| S40 | 25.6 | 248.2 | Detected |
| S41 | 25.7 | 231.5 | Detected |
| S42 | 25.8 | 215.9 | Not Detected |
| S43 | 26.0 | 285.1 | Detected |
| S44 | 26.1 | 264.7 | Detected |
| S45 | 26.3 | 152.4 | Not Detected |
| S46 | 26.6 | 123.7 | Not Detected |
| S47 | 26.6 | 123.7 | Detected |
| S48 | 26.6 | 123.7 | Detected |
| S49 | 26.9 | 100.4 | Detected |
| S50 | 26.9 | 100.4 | Not Detected |
| S51 | 27.3 | 76.0 | Detected |
| S52 | 27.3 | 76.0 | Detected |
| S53 | 27.3 | 76.0 | Detected |
| S54 | 27.4 | 70.9 | Not Detected |
| S55 | 27.5 | 105.3 | Not Detected |
| S56 | 27.6 | 99.8 | Not Detected |
| S57 | 27.7 | 57.5 | Detected |
| S58 | 27.7 | 57.5 | Not Detected |
| S59 | 27.7 | 57.5 | Not Detected |
| S60 | 27.8 | 84.9 | Not Detected |
| S61 | 27.9 | 50.0 | Not Detected |
| S62 | 28.0 | 46.7 | Not Detected |
| S63 | 28.3 | 37.9 | Not Detected |
| S64 | 28.4 | 35.3 | Not Detected |
| S65 | 28.4 | 35.3 | Detected |
| S66 | 28.6 | 30.7 | Not Detected |
| S67 | 28.6 | 30.7 | Not Detected |
| S68 | 28.7 | 46.6 | Detected |
| S69 | 28.7 | 46.6 | Detected |
| S70 | 28.8 | 26.7 | Not Detected |
| S71 | 29.0 | 38.6 | Not Detected |
| S72 | 29.1 | 21.7 | Detected |
| S73 | 29.1 | 35.2 | Not Detected |
| S74 | 29.3 | 18.9 | Detected |
| S75 | 29.7 | 14.3 | Not Detected |
| S76 | 29.7 | 14.3 | Detected |
| S77 | 29.7 | 14.1 | Not Detected |
| S78 | 30.3 | 16.1 | Not Detected |
| S79 | 30.3 | 16.1 | Detected |
| S80 | 30.4 | 8.8 | Not Detected |
| S81 | 30.5 | 13.9 | Not Detected |
| S82 | 30.9 | 6.2 | Not Detected |
| S83 | 31.1 | 9.3 | Not Detected |
| S84 | 31.4 | 7.6 | Not Detected |
| S85 | 31.5 | 4.1 | Detected |
| S86 | 32.3 | 4.2 | Not Detected |
| S87 | 32.4 | 3.9 | Not Detected |
| S88 | 32.8 | 2.9 | Not Detected |
| S89 | 33.1 | 2.4 | Not Detected |
| S90 | 33.1 | 2.4 | Not Detected |
| S91 | 33.4 | 1.9 | Not Detected |

The clinical specimens with low viral copies for testing with SIMA were selected to reach an estimated one RNA copy/mL. The least clinical specimen detected had a Ct value of 31.5 and an estimated concentration of 4.1 RNA copies/mL.

SIMA was further tested with 76 specimens negative for SARS-CoV-2 on the reference assays, comprising 20 VTM specimens and 56 clinical specimens. All 76 specimens tested negative with SIMA.

SIMA did not consistently detect low-copy specimens, as several intervening specimens were undetected. Consequently, the guaranteed limit of detection for SIMA was chosen as 555 RNA copies/mL. Thus, SIMA’s performance characteristics were calculated using positive specimens within this range and the negative Specimens. The MedCalc Diagnostic Test Evaluation Calculator was used, and the parameters are tabulated below.

| Performance Parameter | Value (%) | 95% Confidence Interval |
| --- | --- | --- |
| Sensitivity | 95.0 | 83.1 – 99.4 |
| Specificity | 100.0 | 95.3 – 100.0 |
| Positive Predictive Value | 100.0 | - |
| Negative Predictive Value | 97.4 | 90.8 – 99.8 |
| Accuracy | 98.3 | 93.9 – 99.8 |

### Limit of Detection Results

SIMA’s detection limit was evaluated using a serially diluted high-positive clinical specimen with a BGI ORF1ab assay Ct value of 13 and a Qubit RNA quantification value of 68 ng/µL. The diluted clinical specimen was tested with SIMA in ten replicates. SIMA started detecting the clinical specimens from 10^3^ RNA copies/µL.

| Specimen Concentration / mL | # Replicates tested | # Replicates Detected | ORF1ab (FAM) |  |  | Internal Control (ROX) |  |
| --- | --- | --- | --- | --- | --- | --- | --- |
|  |  |  | Time to ≥700 | Time (Min) | Last Read | Time to ≥700 | Last Read |
| 1.0 | 10 | 7 | 994.0 | 10.6 | 3674.3 | 1385.5 | 734.9 |
| 10.0 | 10 | 10 | 724.0 | 6.1 | 3939.6 | 2102.5 | 694.5 |
| 25.0 | 10 | 10 | 768.5 | 6.8 | 3665.4 | 1744.0 | 613.1 |
| 50.0 | 10 | 10 | 780.0 | 7.0 | 3770.5 | - | 522.5 |
| 100.0 | 10 | 10 | 667.5 | 5.1 | 3790.3 | - | 576.7 |
| 5,000.0 | 10 | 10 | 614.5 | 4.2 | 3749.6 | 1143.0 | 1166.3 |

SARS-CoV-2 RNA was detected at a concentration of 1.0 RNA copies/mL, although this was observed 70% of the time.

### Linearity

To determine the assay linearity, serially diluted specimens were tested in duplicate over a range of 10^3^ to 10^7^.

| Specimen Concentration / $\mu\text{L}$ | SIMA's ORF1ab (FAM) | | | Internal Control (ROX) | |
| --- | --- | --- | --- | --- | --- |
| | Time to $\geq 700$ | Time (Min) | Last Read | Time to $\geq 700$ | Last Read |
| 1,000 | 991 | 10.5 | 3373.4 | - | 614.0 |
| 1,000 | 953 | 9.9 | 3573.9 | - | 597.0 |
| 10,000 | 836 | 7.9 | 3539.7 | - | 593.7 |
| 10,000 | 720 | 6.0 | 3704.5 | - | 506.1 |
| 25,000 | 682 | 5.4 | 3635.9 | - | 601.4 |
| 25,000 | 801 | 7.4 | 3594.4 | - | 599.4 |
| 50,000 | 684 | 5.4 | 3538.5 | - | 627.9 |
| 50,000 | 687 | 5.5 | 3750.4 | - | 568.7 |
| 100,000 | 650 | 4.8 | 3676.4 | - | 484.1 |
| 100,000 | 651 | 4.9 | 3668.1 | - | 576.4 |
| 1,000,000 | 614 | 4.2 | 3743.3 | - | 528.3 |
| 1,000,000 | 615 | 4.3 | 3718.7 | - | 600.0 |
| 10,000,000 | 617 | 4.3 | 3599.2 | 1203 | 1592.4 |
| 10,000,000 | 618 | 4.3 | 3646.0 | 1166 | 1617.9 |

The RNA concentration was plotted against the “Time to rise” using a fluorescence threshold of ≥700. “Time to rise” here is equivalent to Ct values since we do not have thermal cycling in isothermal amplification. A logarithmic trendline was added to forecast their relationship.

An R^2^ statistic greater than 0.7 indicates a very strong correlation between the specimen RNA copies and the “Time to rise” obtained from the SIMA kit, as measured by the fluorescent detection equipment.

### Precision

Both the intra-assay and the inter-assay precision were determined to calculate the precision of SIMA. The Coefficient of Variation (CV; %) was determined for each specimen for the “time to rise” (≥ 700 fluorescence used as threshold) and for the last read fluorescence value.

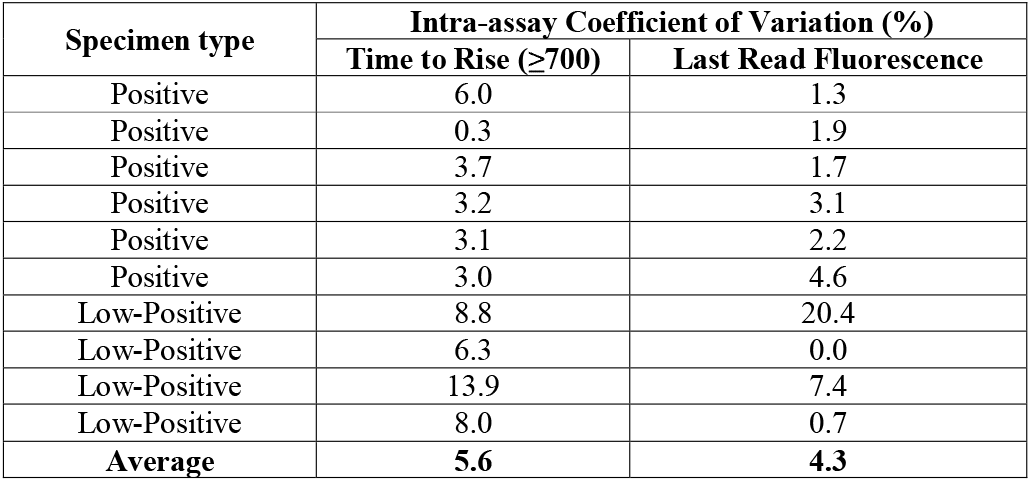

Similarly, the Coefficient of Variation (CV; %) for the inter-assay precision was determined for each specimen for the “time to rise” (≥ 700 fluorescence used as threshold) and for the last read fluorescence value.

| Specimen type | Run Format | Inter-assay Coefficient of Variation (%) |  |
| --- | --- | --- | --- |
| | | Time to rise ($\geq 700$) | Last Read Fluorescence |
| Positive | Once daily over three days | 3.8 | 3.5 |
| Positive | Once daily over three days | 4.3 | 2.8 |
| Low-Positive | Once daily over three days | 10.2 | 20.4 |
| Low-Positive | Once daily over three days | 8.7 | 3.1 |
| Positive | Twice daily, different runs | 1.1 | 2.6 |
| Positive | Twice daily, different runs | 0.8 | 6.5 |
| Low-Positive | Twice daily, different runs | 1.0 | 10.1 |
| **Average** |  | **4.3** | **7.0** |

The intra-assay precision of SIMA was determined to be 5.6 % (Range: 0.3 – 13.9), while its inter-assay precision was 4.3 % (Range: 0.8 – 10.2).

## DISCUSSION AND CONCLUSION

SIMA had a detection limit of 555 RNA copies/µL, with 95% sensitivity and 100% specificity. Its high specificity and POC utility were used to triage workshop participants after the lockdown was lifted. Specimens were collected from participants upon arrival and tested immediately during the registration process. The test results were used to pair participants with their accommodations while isolating the infected individuals in single rooms, allowing them to participate virtually.

SIMA could have a significant public health impact by enabling affordable and timely molecular testing at the point of care in healthcare facilities or at national ports and borders. However, it will not detect new infections with low-viral copies. The protocol for its usage should include retesting after an appropriate interval to rule out new and emerging infections that it may have missed initially.

SIMA design and development were well thought out and extensively validated to demonstrate its fitness for purpose. However, it was not upscaled as projected due to the need for additional detection devices. SIMA solidifies the pathway for the rapid development of diagnostics according to national health needs.

The development of SIMA, a molecular diagnostic assay, was the first in NIMR and Nigeria. This achievement highlights the quality of the staff, facilities, and management at NIMR, which enabled them to rise to the occasion when the nation needed them. More importantly, the development of SIMA was sponsored by a grant from a philanthropic organisation in Nigeria. This grant enabled the development, validation, and launch of the assay within three months, despite a global logistics quagmire. Thus, we can achieve a great deal for Nigeria and the world by collaborating with the well-meaning private sector, companies, and philanthropists to improve health indices.

The way forward will include creating more diagnostic assays using this technology and platform, as it is amenable for use in resource-constrained settings. Having multiple assays available will make the purchase of the detection device economically feasible and positively influence its uptake.

### Limitations

Serially diluted specimens did not consistently detect the human gene used as the internal control, possibly due to an initial low copy number, especially with the use of viral RNA/DNA kits for extraction.

SIMA may not detect atypical specimens, such as those with non-sigmoidal curves or elevated baselines. SIMA did not consistently detect very low-copy specimens, as several intervening specimens were undetected. This could be due to variations in viral genomes and/or the inefficiency of the reverse transcriptase enzyme in reverse transcribing sufficient copies within six minutes at a low temperature. Thus, a guaranteed detection limit has been determined.

However, a negative result by a molecular test does not conclusively rule out the possibility of infection, especially with novel viruses. All results should be interpreted by a qualified healthcare professional, taking into account the patient’s medical history and clinical presentation.

## Data Availability

All data produced in the present work are contained in the manuscript.

## Acknowledgement

We acknowledge the grant support from the Fate Philanthropy Coalition against COVID-19 (FPCC) Support Fund, which enabled the design and development of SIMA amid the COVID-19 pandemic lockdown. FPCC initiated a call with NIMR, asking how to assist NIMR in serving the people. FPCC subsequently selected the SIMA grant out of three proposals received. We appreciate and salute their initiative and partnership.

The Diagnostic Assays Development (DAD) Programme developed SIMA at the Nigerian Institute of Medical Research (NIMR). The DAD programme is an activity of the Population Genomics and Cancer (POPGEC) research group, led by Dr. C.K. Onwuamah, at the Centre for Human Virology and Genomics, Department of Microbiology, NIMR. Thus, SIMA is a product of NIMR’s Research Innovation and Product Development.

